# Use of systemic hormonal contraception and risk of attempted suicide

**DOI:** 10.1101/2022.06.14.22276379

**Authors:** Elena Toffol, Oskari Heikinheimo, Timo Partonen, Anna But, Antti Latvala, Jari Haukka

**Author notes:** **Correspondence to:** Dr. Elena Toffol, Department of Public Health, Clinicum, University of Helsinki, Helsinki, Finland.

## Abstract

**Background:** Evidence regarding the role of hormonal contraception (HC) as a risk factor for attempted suicide (AS) is inconclusive. The aim was to assess the associations of use of different types of systemic hormonal contraceptives with the risk of AS in women aged 15–49 years.

**Methods:** Data were retrieved from national registers in Finland. The AS incidence in 2018-2019 was assessed in a population-based cohort of women (n=587,823) according to their HC use in 2017. To examine the risk of AS related to HC use in the six months preceding the suicide attempt (i.e. current use), we used a nested case-control design with 1:4 ratio (n=4090), and applied multivariable conditional logistic regression models.

**Results:** During the follow-up 1.174,346 million person-years were cumulated and 818 AS cases observed (incidence rate=0.70, 95% CI=0.63–0.83 per 1000 person-years). The incidence rate ratio of HC (n=344 cases) *vs*. no-HC (n=474 cases) was 0.73 (95% CI=0.63–0.83). Use of HC, and specifically of combined hormonal contraceptives was associated with a lower AS risk compared to non-use when controlling for marital status, socioeconomic status, education, chronic diseases and recent delivery (OR=0.55, 95% CI=0.42–0.73), but not when further adjusting for recent psychiatric hospitalizations and current use of psychotropic medications (OR=0.68, 95% CI=0.45–1.02). Current use of progestogen-only preparations was not associated with AS.

**Conclusions:** HC use appeared not associated with an increased AS risk in fertile-aged women. The choice of the best and safest contraceptive option should take individual mental health status, including assessment of suicidal risk, into account.

## Introduction

The global age-standardized suicide rate in 2019 was 9.0 per 100,000 persons (WHO 2021), which corresponds to more than 700,000 suicidal deaths every year. It is estimated that for every adult who dies by suicide, there are more than 20 people who attempt suicide (WHO 2014). Given that a prior suicidal attempt is the major risk factor for a death from suicide (WHO 2014), it is evident that suicidal behavior represents a major public health problem.

Attempted suicides (AS) are two to three times more common in women than in men, especially in the young age group (Baca-Garcia, Perez-Rodriguez, Mann & Oquendo, 2008; Bachmann, 2018). In addition to current or previous psychiatric disorders, many other biological, social or psychological factors contribute to the risk of AS. The possible role of hormonal contraception (HC) as a risk factor for AS has gained particular attention in recent years, especially with respect to young, fertile-aged women (Charlton et al., 2014; Skovlund, Mørch, Kessing, Lange, & Lidegaard, 2018; Edwards et al., 2020). However, results appear inconsistent (Amarasekera, Catalao, Molyneaux, & Fuhr, 2020), and some studies have shown a reduced risk or no associations between the use of contraception and suicidal risk (Vessey, McPherson, Lawless, & Yeates, 1985; Vessey, Villard-Mackintosh, McPherson, & Yeates, 1989; Hannaford et al., 2010; Keyes et al., 2013). For example, one of the first studies to examine the association of oral contraceptive (OC) use with mortality found no difference in mortality by several causes, including suicide, between the ever, current or past users *vs*. never-users in a 12-year follow-up study of over 100,000 US women aged 30–55 years (Colditz, 1994). However, the 36-year follow-up of the same study found higher risk of suicide (hazard ratio, HR 1.41, 95% CI 1.05–1.87) in ever users *vs*. never-users of OCs, with no duration-of-use-related trend, but with increased risk in relation to longer time since last use (Charlton et al., 2014). On the other hand, in a 14-year longitudinal US study of women aged 25–34 years Keyes et al. (2013) found a reduced (by 62%) risk of past-year AS among HC users compared to users of non-hormonal methods.

Different types of HC are known to have different profiles of side effects and adverse events, including mood effects, mostly depending on the types and doses of estrogens and progestogens (Bitzer & Simon, 2011). However, to the best of our knowledge, only few recent studies have examined the associations of different types and doses of HC with the risk of suicidal behavior (Skovlund et al., 2018; Edwards et al., 2020), with partly inconclusive results.

Thus, the aim of this study was to assess the associations of the current use of HC with the risk of AS. Specifically, we examined the risk of AS in relation to different types of systemic hormonal contraceptives in a large cohort of fertile-aged women using recent (2017–2019) high quality registry data from Finland.

## Methods

### Study population and design

This study was part of a larger register-based study of HC use in Finland (Toffol et al., 2020). Briefly, the population was selected, based on the unique personal identification number that is given at birth or at immigration to each person permanently residing in Finland, as inclusive of all fertile-aged women (15–49 years) with at least one redeemed prescription for HC (Anatomical Therapeutic Chemical -ATC-codes: G02B, “contraceptives for topical use”; G03A, “hormonal contraceptives for systemic use”; G03HB, “antiandrogens and estrogens”) in 2017 (n = 294,445) according to the Prescription Centre in the Kanta Services (https://www.kanta.fi/en/what-are-kanta-services). A 1:1 matched (by age and municipality of residence) reference group of women with no redeemed HC prescriptions in 2017 (HC non-users) was also selected. After exclusion of 89 women with a prescription for emergency contraception (ATC code “G03AD”, usually available without prescription in Finland), and of their matched control individuals, a final population of 294,356 HC users and a same-sized reference group were retained for the analyses. Through the Prescription Centre the HC use of all these women was followed-up until the end of 2019. The study was reviewed by the Ethics Committee of the Faculty of Medicine, University of Helsinki (3/2018). Because this is a register-based study, no individual consent is needed.

The follow-up started on 1 January 2018 and ended on 31 December 2019, with a maximum length of follow-up of two years. A first hospitalization or visit to specialized outpatient care (as recorded in the Care Register for Health Care, CRHC) due to attempted suicide (International Classification of Diseases, Tenth Revision, ICD-10 diagnosis “X[6-7]|X8[0-4]”) or a primary health-care contact due to attempted suicide (as recorded in the Register of Primary Health Care visits, RPHC; ICD-10 diagnosis “X[6-7]|X8[0-4]” or International Classification of Primary Care, ICPC, “P77”) were considered as primary endpoint events. Death, emigration from Finland or end of follow-up were defined as censoring events. Individuals with an endpoint event before the start of follow-up (years 2016 and 2017) were excluded from the study (n=889).

The incidence of AS was examined in the cohorts of HC users and non-users (after exclusion of 889 prevalent AS cases, n = 587,823), which altogether included more than half of all fertile-aged women living in Finland in 2017. This cohort was further used as a sampling frame for a nested case-control study, which explored the risk of AS related to current (i.e., in the six months before the event) HC use. For each incident AS case we selected four controls, matched by year of birth.

### The register data and variables

Information on sociodemographic characteristics of all the study members on 31 December 2017 (age, municipality of residence, civil status, socioeconomic status, highest level of education) was retrieved from Statistics Finland. Information about recent deliveries (within the previous two years) was gathered from the Medical Birth Register. Data on special reimbursement rights for chronic diseases (diabetes, multiple sclerosis, epilepsy, severe psychiatric disorders, connective tissue diseases, ulcerative colitis or Crohn’s disease) were obtained from the Social Insurance Institution of Finland. Data on recent (in the past two years) hospitalizations or visits to specialized outpatient care due to psychiatric disorders (ICD-10 codes F00-F99) were gathered from the CRHC. In addition to selection of the initial study population based on redeemed HC prescriptions in 2017, the Prescription Centre was used to gather information on their HC use in the period 2018– 2019. Only ATC codes with at least five individuals in all categories were used in statistical analyses. Moreover, from the Prescription Centre we gathered information on use of psychotropic medications in the 360 days before the event (ATC codes: N05A, antipsychotics; N05B, anxiolytics; N05C, hypnotics and sedatives; N06A, antidepressants; N06B, psychostimulants, N06C psycholeptics and psychoanaleptics in combination). Use of each substance (hormonal contraceptives and psychotropic drugs) was defined as two or more redeemed prescriptions in a 180-day period.

In the incidence study only an indicator of baseline (during year 2017) HC usage (HC use *vs*. no-HC use) was considered. In the nested case-control design, for each HC substance we defined a categorical variable as follows: non-user (no use in the 180 days before the AS event) and current user (use in 1–180 days before the event). The HC methods of interest and available in Finland in 2018 are summarized in Table S1. Contraceptive implants and the levonorgestrel-releasing intrauterine system were excluded, because they can be used for up to three or five years, and are often provided free-of-charge by single communities, thus not necessitating individual prescription (and as such they are not completely covered in the Prescription Centre database). Additionally, levonorgestrel-only containing oral contraceptives were excluded from further analyses because of the small numbers of users and of associated AS cases.

### Statistical analyses

We used Kaplan-Meier curve to describe the incidence rates of AS in relation to HC use in 2017; incidence rate comparison was performed via Poisson regression analysis.

To take into account matching, in the nested case-control design we utilized conditional logistic regression, with group of utilized HC (current use *vs*. non-use) as the main predictor. In addition to a univariate model, we performed Model 1, controlled for marital status, socioeconomic status and education; Model 2, further controlled for chronic diseases (indicator of chronic diseases before start of follow-up) and recent (in the previous six months or in the previous two years) delivery; and Model 3, which is Model 2 further adjusted for recent (in the previous six months or in the previous two years) psychiatric hospitalizations and use of psychotropic medications in the 360 days before the event. Because the case and control groups were matched by year of birth, age was not included as covariate in the models; rather, age-stratified analyses were performed. Sensitivity analyses were conducted by only considering AS cases recorded in the CRHC (i.e., serious AS requiring hospitalization or specialized outpatient care). Additionally, in order to take into account previous severe psychiatric disorders, the analyses were repeated after exclusion of women with special reimbursement rights for severe psychiatric disorders. Moreover, analyses were also conducted in groups stratified by psychiatric history (those with and without a recent hospitalization due to psychiatric disorders, use of psychotropic drugs or reimbursement rights for severe psychiatric disorders at baseline).

All the analyses were performed with R software version 4.0.5 (R Core Team, 2021).

## Results

During the follow-up 1,174,346 person-years were cumulated and 818 AS cases observed, with an overall incidence rate (IR) of 0.70 (95% CI 0.63–0.83) per 1000 person-years. In the group of HC non-users in 2017, we observed 474 cases of AS (IR 0.81 per 1000 person-years, 95% CI 0.74– 0.88), while among HC users there were 344 AS cases (IR 0.59, 95% CI 0.53–0.65). The incidence rate ratio (IRR) of HC *vs*. no-HC users was 0.73 (0.63–0.83) (Figure 1). The incidence of AS decreased with age, with the highest IR in the age group 15–19 years (1.62, 95% CI 1.42–1.83) (Table S2).

**Figure 1.**
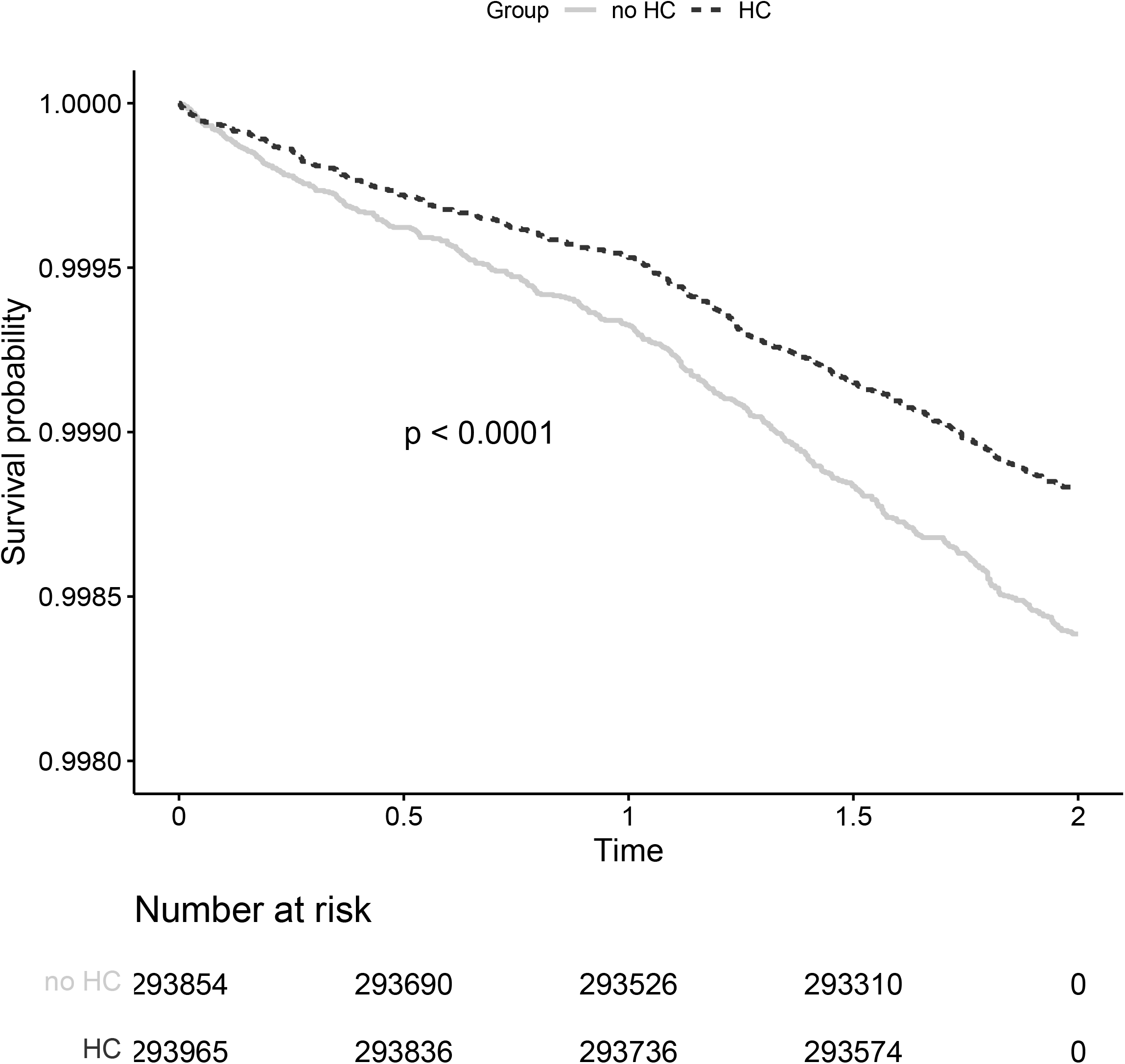
Kaplan-Meier curve for incidence of attempted suicide. *p*-value of log-rank test shown.

The nested case-control study for AS built on this cohort consisted of altogether 4090 women (Table 1). Compared to the controls, women who attempted suicide were less likely to be married, employed, with post-secondary or higher education, and to have given birth in the previous two years, but more likely to have had a recent psychiatric hospitalization. Additionally, they were less likely to be current users of HC (15.6% *vs*. 22.2%), in particular of combined hormonal contraceptives (CHCs) (9.8% *vs*. 17.8%: ethinyl estradiol (EE)-containing preparations, 7.5% *vs*. 13.9%; estradiol-containing preparations, 2.3% *vs*. 4.0%; *p* < 0.001). Specifically, current use of desogestrel and EE (0.9% *vs*. 2.0%, *p* = 0.045) and of drospirenone and EE (3.7% *vs*. 6.3%, *p* = 0.005) was less common among women who attempted suicide than in their controls (Table 2). Consistently, in the univariable logistic regression model, use of HC (OR 0.64, 95% CI 0.52–0.79), and specifically of CHCs (either EE-containing or estradiol-containing preparations) was associated with lower risk of AS compared to the risk in HC non-users (OR 0.50, 95% CI 0.39–0.64). No significant associations emerged with current use of progestin-only contraceptives (Table 3). The lower risk associated with CHCs (specifically, EE-containing contraceptives) remained significant after controlling for covariates (Models 1–2), except when further adjusting for recent psychiatric hospitalizations and current use of psychotropic medications (Model 3, see Table 3). The results did not change in age-stratified analyses (Table S3). In analyses stratified by psychiatric history, current use of HC, and in detail of EE-containing preparations, was associated with lower risk of AS only in women without psychiatric disorders (Table S4). Among women with psychiatric disorders HC use was not associated with AS risk.

**Table 1.** Basic characteristics of the nested case-control study of attempted suicides.

|  | **Cases**<br>**(N=818)** |  | **Controls**<br>**(N=3272)** |  |  |
| --- | --- | --- | --- | --- | --- |
|  | **N** | **%** | **N** | **%** | **p-value** |
| **Marital status** |  |  |  |  | <0.001 |
| Unmarried | 667 | 81.5 | 2643 | 80.8 |  |
| Married | 84 | 10.3 | 545 | 16.7 |  |
| Divorced | 65 | 7.9 | 79 | 2.4 |  |
| Widowed | <5 | NA | <5 | NA |  |
| Other | <5 | NA | <5 | NA |  |
| **Socioeconomic group** |  |  |  |  | <0.001 |
| Self-employed | 19 | 2.3 | 96 | 3.0 |  |
| Upper-level employees | 22 | 2.7 | 308 | 9.5 |  |
| Lower-level employees | 118 | 14.5 | 850 | 26.2 |  |
| Manual workers | 112 | 13.8 | 482 | 14.9 |  |
| Students | 281 | 34.6 | 1067 | 32.9 |  |
| Pensioners | 70 | 8.6 | 41 | 1.3 |  |
| Others | 122 | 15.0 | 216 | 6.7 |  |
| Unknown | 69 | 8.5 | 185 | 5.7 |  |
| **Education** |  |  |  |  | <0.001 |
| Upper secondary | 325 | 39.7 | 1488 | 45.5 |  |
| Post-secondary non-tertiary | <5 | NA | 15 | 0.5 |  |
| Short-cycle tertiary | 6 | 0.7 | 39 | 1.2 |  |
| Bachelor | 29 | 3.5 | 471 | 14.4 |  |
| Master | 17 | 2.1 | 190 | 5.8 |  |
| Doctoral | <5 | NA | 12 | 0.4 |  |
| Missing (including, e.g., missing information on education other than of primary school level, school dropouts) | 439 | 53.7 | 1057 | 32.3 |  |
| **Age group<sup>\*</sup>** |  |  |  |  | 1.000 |
| 15-19 years | 249 | 30.4 | 996 | 30.4 |  |
| 20-24 years | 239 | 29.2 | 956 | 29.2 |  |
| 25-29 years | 143 | 17.5 | 572 | 17.5 |  |
| 30-34 years | 79 | 9.7 | 316 | 9.7 |  |
| 35-39 years | 56 | 6.8 | 224 | 6.8 |  |
| 40-44 years | 35 | 4.3 | 140 | 4.3 |  |
| 45-49 years | 17 | 2.1 | 68 | 2.1 |  |
| **Severe psychiatric disorder at baseline** | 104 | 12.7 | 24 | 0.7 | <0.001 |
| **Chronic somatic disease at baseline\*\*** | 60 | 7.3 | 146 | 4.5 | 0.001 |
| **HC use in 2017** | 344 | 42.1 | 1668 | 51.0 | <0.001 |
| **Use of psychotropic medication** |  |  |  |  |  |
| antipsychotics | 281 | 34.4 | 59 | 1.8 | <0.001 |
| anxiolytics | 186 | 22.7 | 31 | 0.9 | <0.001 |
| hypnotics and sedatives | 112 | 13.7 | 41 | 1.3 | <0.001 |
| antidepressants | 399 | 48.8 | 240 | 7.3 | <0.001 |
| psychostimulants | 13 | 1.6 | 13 | 0.4 | <0.001 |
| psycholeptics and psychoanaleptics in combination | 5 | 0.6 | <5 | NA | 0.001 |
| **Recent delivery** |  |  |  |  | <0.001 |
| No | 799 | 97.7 | 3077 | 94.0 |  |
| In the previous 6 months | < 5 | NA | 29 | 0.9 |  |
| 6 to 24 months before | 17 | 2.1 | 166 | 5.1 |  |
| **Recent hospitalization due to psychiatric disorders** |  |  |  |  | <0.001 |
| No | 189 | 23.1 | 2978 | 91.0 |  |
| In the previous 6 months | 524 | 64.1 | 175 | 5.3 |  |
| 6 to 24 months before | 105 | 12.8 | 119 | 3.6 |  |

\* matched by age
\*\* Diabetes, multiple sclerosis, epilepsy, connective tissue diseases, ulcerative colitis or Chron's disease.
not
available

**Table 2.** Hormonal contraception use in the nested case-control study of attempted suicides.

|  | **Cases**<br>**(N=818)** |  | **Controls**<br>**(N=3272)** |  |  |
| --- | --- | --- | --- | --- | --- |
|  | **N** | **%** | **N** | **%** | **p-value** |
| **HC group** |  |  |  |  | <0.001 |
| No HC | 690 | 84.4 | 2545 | 77.8 |  |
| Current HC | 128 | 15.6 | 727 | 22.2 |  |
| Combined hormonal contraception | 80 | 9.8 | 584 | 17.8 |  |
| Ethinyl-estradiol containing | 61 | 7.5 | 454 | 13.9 |  |
| Estradiol containing | 19 | 2.3 | 130 | 4.0 |  |
| Progestin-only | 48 | 5.9 | 143 | 4.4 |  |
| **Combined hormonal contraceptives (ATC code)** |  |  |  |  |  |
| Levonorgestrel and ethinylestradiol (G03AA07) | <5 | NA | 14 | 0.4 | 0.124 |
| Desogestrel and ethinylestradiol (G03AA09) | 7 | 0.9 | 64 | 2.0 | 0.045 |
| Gestodene and ethinylestradiol (G03AA10) | 14 | 1.7 | 85 | 2.6 | 0.178 |
| Drospirenone and ethinylestradiol (G03AA12) | 30 | 3.7 | 206 | 6.3 | 0.005 |
| Norelgestromin and ethinylestradiol patch (G03AA13) | <5 | NA | 10 | 0.3 | 0.597 |
| Nomegestrol and estradiol (G03AA14) | <5 | NA | 39 | 1.2 | 0.116 |
| Dienogest and ethinylestradiol (G03AA16) | <5 | NA | 18 | 0.6 | 0.186 |
| Dienogest and estradiol-valerate (G03AB08) | <5 | NA | 27 | 0.8 | 0.052 |
| Etonogestrel and ethinylestradiol vaginal ring (G02BB01) | 8 | 1.0 | 58 | 1.8 | 0.145 |
| **Progestin-only oral contraceptives** |  |  |  |  |  |
| Norethisterone (G03AC01) | 5 | 0.6 | 8 | 0.2 | 0.187 |
| Desogestrel (G03AC09) | 42 | 5.1 | 135 | 4.1 | 0.241 |
| **Antiandrogen and estrogen** |  |  |  |  |  |
| Cyproterone and estrogen (G03HB01) | 14 | 1.7 | 65 | 2.0 | 0.712 |

ATC, Anatomical Therapeutic Chemical; NA, Not Available.

**Table 3.** Nested case-control study of AS. Odds ratios and 95% confidence intervals based on conditional logistic regression models. Usage of drugs in respect to the event day: no=no use in the past 180 days; current= use in the past 180 days. Multivariate Model 1 is adjusted with the following covariates: marital status, socioeconomic status, education; Model 2 is Model 1 further adjusted for chronic diseases (reimbursement codes^*^) and recent delivery; Model 3 is Model 2 further adjusted for recent psychiatric hospitalization and current use of psychotropic medications.

|  |  | Univariable |  | Model 1 |  | Model 2 |  | Model 3 |  |
| --- | --- | --- | --- | --- | --- | --- | --- | --- | --- |
|  |  | Odds Ratio | 95% CI | Odds Ratio | 95% CI | Odds Ratio | 95% CI | Odds Ratio | 95% CI |
| **CRHC and RPHC data** |  |  |  |  |  |  |  |  |  |
| **HC groups** | No HC | reference |  | reference |  | reference |  | reference |  |
|  | Current HC | 0.64 | 0.52–0.79 | 0.74 | 0.59–0.93 | 0.71 | 0.56–0.90 | 0.79 | 0.56–1.11 |
|  | CHC | 0.50 | 0.39–0.64 | 0.59 | 0.45–0.77 | 0.55 | 0.42–0.73 | 0.68 | 0.45–1.02 |
|  | EE containing CHCs | 0.49 | 0.37–0.64 | 0.56 | 0.41–0.75 | 0.54 | 0.40–0.73 | 0.74 | 0.47–1.17 |
|  | estradiol containing CHCs | 0.53 | 0.33–0.87 | 0.71 | 0.43–1.20 | 0.60 | 0.34–1.05 | 0.50 | 0.21–1.19 |
|  | Progestin-only | 1.24 | 0.88–1.74 | 1.40 | 0.96–2.06 | 1.39 | 0.94–2.05 | 1.08 | 0.62–1.88 |
| **Only CRHC data** |  |  |  |  |  |  |  |  |  |
| **HC groups** | No HC | reference |  | reference |  | reference |  | reference |  |
|  | Current HC | 0.59 | 0.48–0.73 | 0.65 | 0.51–0.82 | 0.65 | 0.51–0.82 | 0.86 | 0.61–1.21 |
|  | CHC | 0.46 | 0.35–0.60 | 0.49 | 0.37–0.65 | 0.49 | 0.37–0.65 | 0.72 | 0.48–1.08 |
|  | EE containing CHCs | 0.46 | 0.34–0.62 | 0.49 | 0.36–0.68 | 0.50 | 0.36–0.69 | 0.78 | 0.50–1.24 |
|  | estradiol containing CHCs | 0.46 | 0.28–0.75 | 0.50 | 0.30–0.85 | 0.46 | 0.27–0.81 | 0.56 | 0.26–1.24 |
|  | Progestin-only | 1.08 | 0.77–1.54 | 1.29 | 0.88–1.90 | 1.36 | 0.91–2.02 | 1.35 | 0.74–2.46 |

<sup>\*</sup>diabetes, multiple sclerosis, epilepsy, severe psychiatric disorders, connective tissue diseases, ulcerative colitis or Crohn's disease
CHC, combined hormonal contraception; CRHC, Care Register for Health Care; EE, ethinyl estradiol; HC, hormonal contraception; RPHC, Register of Primary Health Care visit.

In detail, in unadjusted models current use of combined OCs (COC) containing desogestrel and EE, and drospirenone and EE was associated with lower AS risk than non-use of the same preparations (OR 0.43, 95% CI 0.20–0.94; and OR 0.56, 95% CI 0.38–0.84, respectively). The association with drospirenone and EE remained significant after controlling for background characteristics (socioeconomic status, marital status, education, chronic diseases, recent delivery) and after exclusion of women with previous severe psychiatric disorders, but was lost when further adjusting for recent psychiatric hospitalizations and use of psychotropic medications (Figure 2).

**Figure 2.**
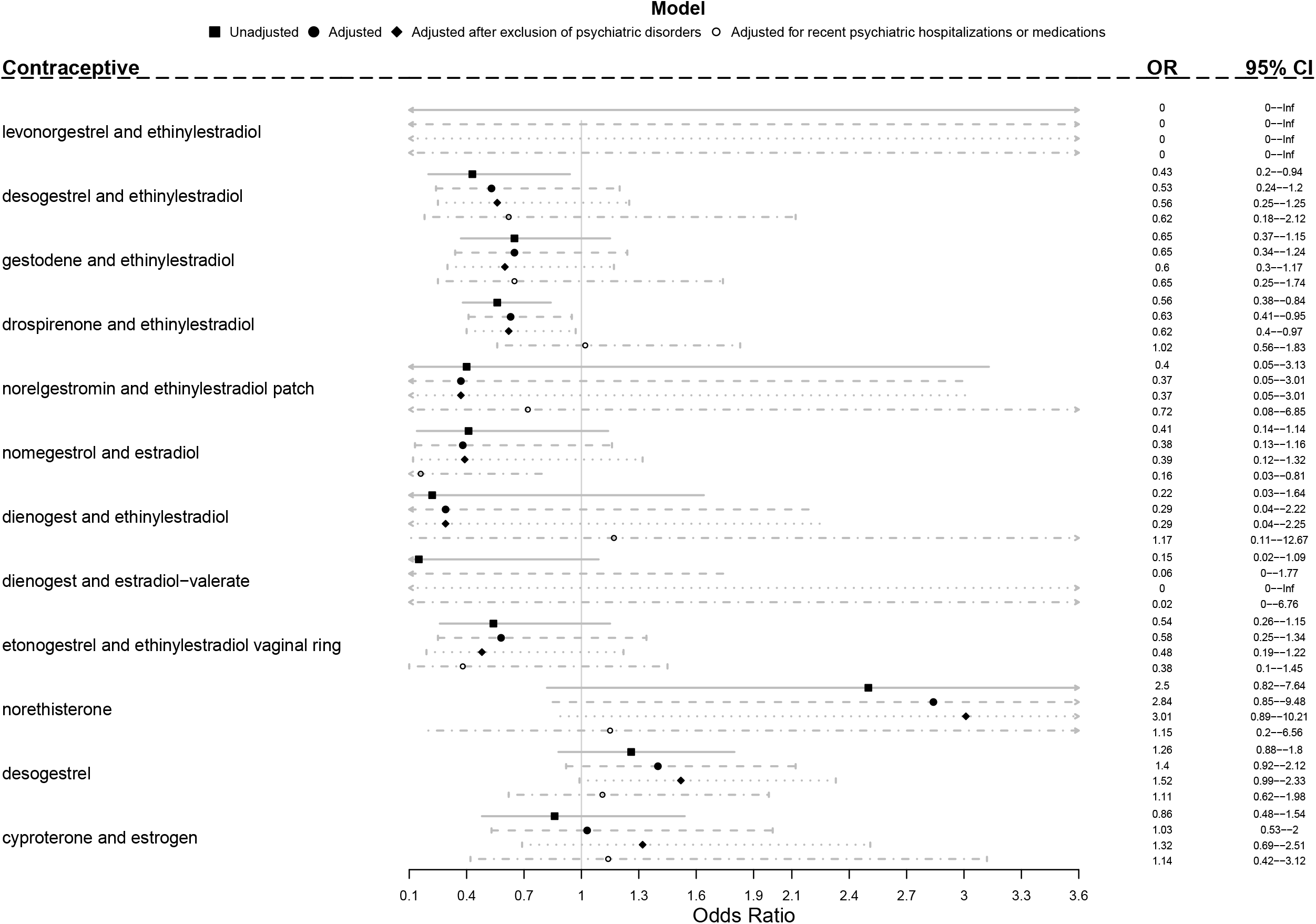
Associations between attempted suicide and current use (in the previous 180 days) of hormonal contraceptives. Results are expressed as Odds Ratios with 95% Confidence Intervals. For each substance the reference category is no use of the same substance in the 180 days before the attempted suicide. Adjusted model is controlled for marital status, socioeconomic status, education, chronic diseases and recent delivery. One substance in model a time. Care Register of Health Care and Register of Primary Health Care visits data together.

In sensitivity analyses including only AS cases recorded in the CRHC, among the 1,174,475 person-years cumulated 760 AS cases were observed (IR 0.65 per 1000 person-years, 95% CI 0.60– 0.70). Of them, 440 occurred among HC non-users in 2017, and 320 in HC users, with an IRR of HC *vs*. no-HC users of 0.73 (95% CI 0.63–0.84; see Table S5). Basic characteristics of the population of the nested-case control study based on CRHC cases only are reported in Table S6 and Table S7. Associations with AS risk were similar to those in the original population (Table 3, Figure S1). In addition, current use of norethisterone-only OC was associated with higher AS risk when controlling for background characteristics (OR 4.78, 95% CI 1.34–21.72) and after exclusion of women with previous severe psychiatric disorders (OR 5.63, 95% CI 1.46–21.73), but not when adjusting for recent psychiatric hospitalizations and use of psychotropic medications (OR 1.40, 95% CI 0.20–9.63) (Figure S1).

## Discussion

The main finding of our study is that HC use was not associated with an increased risk of attempted suicide in women of fertile age. In addition, the current use of CHCs, and in particular of EE-containing preparations was associated with a lower risk of attempted suicide compared to non-use of HC even after controlling for covariates and in women without psychiatric disorders. However, the significance of this finding was lost after taking psychiatric disorders into account.

Differently from suicide, epidemiological data on AS are difficult to retrieve and often inconsistent, due to the lack of official registration systems and different definitions of suicidal attempts. Subsequently, the overall incidence of AS in our population (0.70 per 1000 women-years) corresponds only in part to previously reported figures (Bossuyt & Van Casteren, 2007). For example, a previous register-based study from Finland reported an incidence of first serious AS in men and women aged 12 or older of 0.4 per 1000 person-years between 1996 and 2003 (Haukka, Suominen, Partonen, & Lönnqvist, 2008). On the other hand, a prospective study conducted between 1981 and 1993 in a community sample in Baltimore found an incidence of 1.6 per 1000 women-years in women of all ages (specifically, 3.7 per 1000 women-years in the age group 18–29 years, and 1.2 per 1000 women-years in the age group 30–44 years) (Kuo, Gallo, & Tien, 2001). Similarly, in a recent Danish study (Skovlund et al., 2018) the incidence of AS was 1.8 per 1000 women-years in women aged 15–33 years (compared to an IR of 0.8 per 1000 women-years in the same age group, i.e. 15–34 years, in our study). Differences between the above reported figures are attributable, among others, to different times of assessment, with consequent different levels of awareness and preventive measures implemented to contrast suicidal behavior, and to different social and cultural milieu affecting contraceptive choices. Additionally, the use of interview-reported cases, thus including AS which did not receive medical attention, likely explains the higher AS rates reported in some studies (Kuo et al., 2001).

Our finding that the current use of HC is not associated with an increased risk of AS in fertile-aged women contrasts with results reported by recent studies in the Nordic countries. A Danish study found a higher risk of the first AS in women aged 15–33 years using HC both currently (relative risk, RR 1.97, 95% CI 1.85–2.10) and previously (RR 3.40, 95% CI 3.11–3.71) compared with never-users (Skovlund et al., 2018). Similarly, a large Swedish register-based study of women aged 15–22 years found an increased risk of suicidal behavior (attempted or completed suicide) in users of COCs (HR 1.36, 95%CI 1.18–1.56) and progestin-only pills (HR 1.75, 95% CI 1.44–2.12) (Edwards et al., 2020). A Korean study also reported positive associations between OC use and suicidality (ideation and attempt) in women over 20 years of age; however, the latter compared, with a retrospective design, lifetime prevalence of OC use and period prevalence of suicidality, and did not provide any information on the type of OC or on other types of HC (Jung, Cho, & Kim, 2019). One of the possible reasons underlying these different findings is the inclusion in our sample of older women, where the prevalence of AS in general is lower, and who usually have a distinct profile of HC use. On this regard, it must be acknowledged that older women are more likely to use long-active reversible contraceptive (LARC) methods, such as the levonorgestrel-releasing intrauterine system, which were not included in our study, given that most of these methods are provided free-of-charge from some municipalities in Finland, and can be used for up to five years. However, our results did not change in age-stratified analyses. Another possible explanation to our findings is that women with severe psychiatric disorders may be less likely to use HC (Toffol et al., 2020), but more likely to attempt suicide. However, no associations between HC use and higher risk of AS emerged in analyses stratified by psychiatric history.

The observation of a negative association between current use of EE-containing preparations and AS risk is a novel one, although the association lost its significance after controlling for psychiatric disorders, which are known to be the main risk factor for suicidal behavior. Skovlund et al. (2018) found an increased risk associated with the use of all EE-containing COCs, but the highest risk was seen in users of patch, vaginal ring and progestin-only products. While the Danish study took the use of antidepressants or psychiatric diagnoses after starting the HC into account as a possible mediator factor in the relationship between HC use and suicidal behavior, we controlled our results for the current use of any psychotropic medications, including antipsychotics and anxiolytics. This may be relevant, as we have previously shown positive associations between HC use and the use of psychotropic drugs of any class in the same population as in the current cohort used to assess incidence (Toffol et al., 2022). We also found a tendency for a higher AS risk with the use of progestin-only products, such as norethisterone and desogestrel containing oral contraceptives, but no associations with the use of vaginal ring and patch. Even though the negative impact of norethisterone on mood has been previously observed (Lawrie, Hofmeyr, De Jager, Berk, Paiker, & Viljoen, 1998; Dennis, Ross, & Herxheimer, 2008), our results have to be taken with caution, given that the use of norethisterone-only pills in Finland is rather limited (Table S1), especially to the postpartum period. However, the tendency did not change after controlling for a recent delivery.

Another key finding of our study is a tendency for a lower AS risk in women using drospirenone and EE. Drospirenone has progestogenic, antimineralcorticoid and antiandrogenic activities. These characteristics are related to its favorable influence on mood also in women suffering from premenstrual dysphoric disorder (Paoletti et al., 2004; Pearlstein, Bachmann, Zacur, & Yonkers, 2005; Sangthawan, & Taneepanichskul, 2005; Yonkers et al., 2005). This finding is of relevance, as OC containing drospirenone (and third generation progestins such as desogestrel, etonogestrel and gestodene) are used more commonly in Finland than in the other Nordic countries, and OC containing drospirenone is the most used COC in Finland (Table S1).

This study has a number of limitations. By using registry data, we were not able to detect less severe cases of AS which remained unknown to the health-care system, which likely account for almost half of the self-reported AS (Kjøller, & Helweg-Larsen, 2000). Additionally, as HC use was defined based on redeemed prescriptions rather than on its actual use, misclassification cannot be ruled out. However, because the Social Insurance Institution in Finland does not reimburse HC, it is likely that the majority of those who purchased the drug did in fact use it. Likewise, it cannot be excluded that women currently not using HC at the time of the AS (in the 6 months before) were in fact former users, who discontinued their contraceptive due to side effects, including mood changes. For example, Skovlud et al. (2018) found a higher risk of AS in former users than in current users of HC. However, in our nested case-control study, most of women who attempted suicide in the period 2018–2019 were in fact HC non-users in 2017 (57.9% *vs*. 49.0% in the control group). Another limitation arises from the lack of information on the precise contents of the contraceptive preparations used, which precluded any analyses on the effect of different doses of EE. Additionally, we lacked information on the use of non-hormonal methods (e.g., copper intrauterine device and barrier methods) as well as contraceptives obtained free-of-charge as part of municipal programs, especially LARC methods. Furthermore, we cannot exclude that the detected associations are confounded by external unaccounted factors.

Among the strengths of our study is the use of Finnish register data of proven high quality (Heino, Niinimäki, Mentula, & Gissler, 2018; Sund, 2012), and the identification of AS cases based on diagnostic codes from specialist health-care between 2018 and 2019. The nested case-control design is shown to produce unbiased estimates and to be free from weaknesses of ordinary case-control design. It uses correct sampling of controls that takes the follow-up time into account (Langholz, & Richardson, 2009; Sedgwick, 2014). In addition, the control women were matched by age. Thus, our study design provides results that are relatively free from confounding bias, although some residual confounding is always possible in observational studies. Importantly, our results appeared consistent across the two different study designs (incidence study, nested case-control design) and definitions of HC use (baseline and current use).

Taken together, our results convey the reassuring message to fertile-aged women seeking contraception that HC use is not associated with an increased risk of attempted suicide. At the same time, they once more stress the importance of a personalized choice of the best and safest contraceptive option, which should take individual mental health status including assessment of possible suicidal risk into account.

## Supporting information

Heading Supplemental Figure 1

## Data Availability

The data that support the findings of this study are available from Statistics Finland, the Finnish Institute for Health and Welfare, and the Social Insurance Institution, but restrictions apply to the availability of these data, which were used under license for the current study, and so are not publicly available. Data are, however, available from the authors upon reasonable request and with permission of FinData (https://www.findata.fi/en/).

