## Supplementary figures and images for "Use of systemic hormonal contraception and risk of attempted suicide"

### Supplemental Figure 1

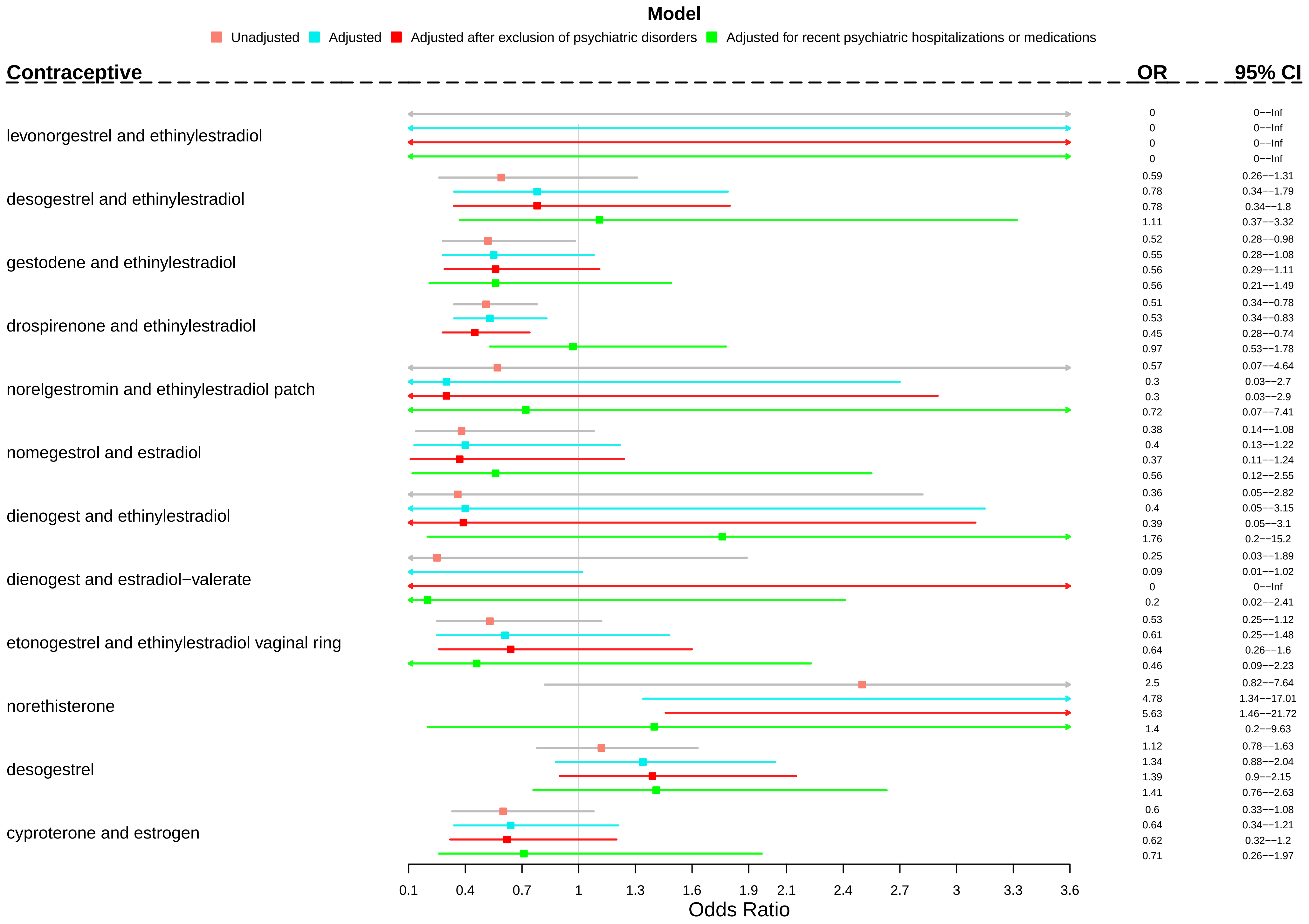
