## Supplemental Tables for "Use of systemic hormonal contraception and risk of attempted suicide"

**Table S1. Systemic hormonal contraceptives, the doses available and their use in Finland in 2018 (https://www.fimea.fi/web/en/databases_and_registers/fimeaweb).**

| **ATC code** | **Available doses** | | **Use in 2018^*^** |
| --- | --- | --- | --- |
| **Combined hormonal contraceptives** | **Monophasic** | **Phasic** |  |
| G03AA07 (levonorgestrel and ethinylestradiol) | 150 μg and 30 μg | 50-125 μg and 30-40 μg | 0.68 |
| G03AA09 (desogestrel and ethinylestradiol) | 150 μg and 20/30 μg | 25-125 μg and 30-40 μg | 3.44 |
| G03AA10 (gestodene and ethinylestradiol) | 75 μg and 20/30 μg | 20-100 μg and 30-40 μg | 5.13 |
| G03AA11 (norgestimate and ethinylestradiol) | 250 μg and 35 μg |  | 0.04 |
| G03AA12 (drospirenone and ethinylestradiol) | 3 mg and 20/30 μg |  | 11.3 |
| G03AA13 (norelgestromin and ethinylestradiol patch) | 6 mg and 600 μg |  | 0.48 |
| G03AA14 (nomegestrol and estradiol) | 2.5 mg and 1.5 mg |  | 3.13 |
| G03AA16 (dienogest and ethinylestradiol) | 2 mg and 30 μg |  | 0.47 |
| G03AB08 (dienogest and estradiol-valerate) |  | 2-3 mg and 1-3 mg | 1.36 |
| G02BB01 (etonogestrel and ethinylestradiol vaginal ring) | 120 μg and 15 μg |  | 2.86 |
| **Progestin-only oral contraceptives** |  |  |  |
| G03AC01 (norethisterone) | 350 μg |  | 0.12 |
| G03AC03 (levonorgestrel) | 30 μg |  | 0.45 |
| G03AC09 (desogestrel) | 75 μg |  | 10.93 |
| **Antiandrogen and estrogen** |  |  |  |
| G03HB01 (cyproterone and estrogen) | 2 mg and 35 μg (EE) | 1-2 mg and 1-2 mg (E2) | 3.34 |

^*^ Defined daily doses (DDD) / 1000 inhabitants/day

**Table S2. Attempted Suicide incidence.** Incidence rate ratio (IRR) with 95% confidence interval based on Poisson regression model.

|  |  | **Person-years** | **Events** | **Rate (1/1000)** | **95% CI** | **IRR** | **95% CI** |
| --- | --- | --- | --- | --- | --- | --- | --- |
| **HC use in year 2017** | no HC | 586.962 | 474 | 0.81 | 0.74, 0.88 | reference | reference |
|  | HC | 587.384 | 344 | 0.59 | 0.53, 0.65 | 0.73 | 0.63, 0.83 |
| **Age group** | 15-19 years | 154.090 | 249 | 1.62 | 1.42, 1.83 | reference | reference |
|  | 20-24 years | 281.188 | 239 | 0.85 | 0.75, 0.97 | 0.53 | 0.44, 0.63 |
|  | 25-29 years | 254.721 | 143 | 0.56 | 0.47, 0.66 | 0.35 | 0.28, 0.43 |
|  | 30-34 years | 176.566 | 79 | 0.45 | 0.35, 0.56 | 0.28 | 0.22, 0.36 |
|  | 35-39 years | 134.366 | 56 | 0.42 | 0.32, 0.54 | 0.26 | 0.19, 0.35 |
|  | 40-44 years | 100.540 | 35 | 0.35 | 0.24, 0.48 | 0.22 | 0.15, 0.31 |
|  | 45-49 years | 72.876 | 17 | 0.23 | 0.14, 0.37 | 0.14 | 0.09, 0.24 |
| **Socioeconomic group** | Self-employed | 41.495 | 19 | 0.46 | 0.28, 0.72 | reference | reference |
|  | Upper-level employees | 143.866 | 22 | 0.15 | 0.10, 0.23 | 0.33 | 0.18, 0.62 |
|  | Lower-level employees | 318.341 | 118 | 0.31 | 0.26, 0.37 | 0.68 | 0.42, 1.10 |
|  | Manual workers | 174.378 | 112 | 0.64 | 0.53, 0.77 | 1.40 | 0.86, 2.28 |
|  | Students | 240.112 | 281 | 1.17 | 1.04, 1.32 | 2.56 | 1.61, 4.07 |
|  | Pensioners | 20.517 | 70 | 3.41 | 2.66, 4.31 | 7.45 | 4.49, 12.37 |
|  | Others | 101.717 | 122 | 1.20 | 1.00, 1.43 | 2.62 | 1.62, 4.25 |
|  | Unknown | 60.891 | 69 | 1.13 | 0.88, 1.43 | 2.48 | 1.49, 4.11 |
| **Education** | Upper secondary | 544.112 | 325 | 0.60 | 0.53, 0.67 | reference | reference |
|  | Post-secondary non-tertiary | 7.483 | <5 | 1.13 | 0.00, 0.75 | 0.22 | 0.03, 1.59 |
|  | Short-cycle tertiary | 30.799 | 6 | 0.20 | 0.07, 0.42 | 0.33 | 0.15, 0.73 |
|  | Bachelor | 224.229 | 29 | 0.13 | 0.09, 0.19 | 0.22 | 0.15, 0.32 |
|  | Master | 124.131 | 17 | 0.14 | 0.08, 0.22 | 0.23 | 0.14, 0.37 |
|  | Doctoral | 6.529 | <5 | 0.15 | 0.004, 0.85 | 0.26 | 0.04, 1.83 |
|  | Unknown | 237.063 | 439 | 1.85 | 1.68, 2.03 | 3.10 | 2.69, 3.58 |
| **Marital status** | Unmarried | 805.364 | 667 | 0.83 | 0.77, 0.89 | reference | reference |
|  | Married | 302.026 | 84 | 0.28 | 0.22, 0.34 | 0.34 | 0.27, 0.42 |
|  | Divorced | 63.214 | 65 | 1.03 | 0.79, 1.31 | 1.24 | 0.96, 1.60 |
|  | Widowed | 2.184 | <5 | 0.46 | 0.01, 2.55 | 0.55 | 0.08, 3.93 |
|  | Other | 1.558 | <5 | 0.64 | 0.02,-3.58 | 0.78 | 0.11, 5.51 |

HC, hormonal contraception

**Table S3. Associations between current HC use and attempted suicide in age-stratified analyses.** Model controlled for marital status, socioeconomic status, education, chronic diseases (reimbursement codes^*^), recent delivery, recent psychiatric hospitalization and current use of psychotropic medications.

|  | | **15-19 years** | | **20-24 years** | | **25-34 years** | | **35-49 years** | |
| --- | --- | --- | --- | --- | --- | --- | --- | --- | --- |
|  |  | **Odds Ratio** | **95% CI** | **Odds Ratio** | **95% CI** | **Odds Ratio** | **95% CI** | **Odds Ratio** | **95% CI** |
| **HC groups** | No HC | reference | | reference | | reference | | reference | |
|  | Current HC | 0.96 | 0.55–1.67 | 0.65 | 0.35–1.22 | 0.53 | 0.24–1.20 | 0.69 | 0.15–3.14 |
|  | CHC | 0.85 | 0.45–1.63 | 0.57 | 0.28–1.17 | 0.44 | 0.17–1.17 | 2.85 | 0.20–40.96 |
|  | Progestin-only | 1.33 | 0.48–3.73 | 0.95 | 0.31–2.88 | 0.80 | 0.20–3.13 | 0.42 | 0.07–2.46 |

^*^diabetes, multiple sclerosis, epilepsy, severe psychiatric disorders, connective tissue diseases, ulcerative colitis or Crohn’s disease

CHC, Combined hormonal contraception; HC, hormonal contraception

**Table S4. Associations between current HC use and attempted suicide in analyses stratified by psychiatric history.** Model controlled for marital status, socioeconomic status, education, chronic somatic diseases^*^ and recent delivery.

|  | | **No psychiatric history** | | **Psychiatric history** | |
| --- | --- | --- | --- | --- | --- |
|  |  | **Odds Ratio** | **95% CI** | **Odds Ratio** | **95% CI** |
| **HC groups** | No HC | reference | | reference | |
|  | Current HC | 0.73 | 0.58–0.91 | 0.78 | 0.50–1.21 |
|  | CHC | 0.57 | 0.44–0.75 | 0.64 | 0.37–1.11 |
|  | EE containing CHCs | 0.54 | 0.40–0.73 | 0.63 | 0.33–1.19 |
|  | estradiol containing CHCs | 0.69 | 0.41–1.16 | 0.68 | 0.25–1.84 |
|  | Progestin-only | 1.41 | 0.96–2.06 | 1.10 | 0.54–2.23 |

^*^diabetes, multiple sclerosis, epilepsy, connective tissue diseases, ulcerative colitis or Crohn’s disease

CHC, Combined hormonal contraception; EE, ethinyl estradiol; HC, hormonal contraception

**Table S5. Attempted suicide incidence.** Incidence rate ratio (IRR) with 95% confidence interval based on Poisson regression model. Only cases from Care Register for Health Care included.

|  |  | **Person-years** | **Events** | **Rate (1/1000)** | **95% CI** | **IRR** | **95% CI** |
| --- | --- | --- | --- | --- | --- | --- | --- |
| **HC use in year 2017** | no HC | 587.040 | 440 | 0.75 | 0.68, 0.82 | reference | reference |
|  | HC | 587.435 | 320 | 0.55 | 0.49, 0.61 | 0.73 | 0.63, 0.84 |
| **Age group** | 15-19 years | 154.135 | 229 | 1.49 | 1.30, 1.69 | reference | reference |
|  | 20-24 years | 281.227 | 224 | 0.80 | 0.70, 0.91 | 0.54 | 0.45, 0.65 |
|  | 25-29 years | 254.742 | 135 | 0.53 | 0.44, 0.63 | 0.36 | 0.29, 0.44 |
|  | 30-34 years | 176.575 | 76 | 0.43 | 0.34, 0.54 | 0.29 | 0.22, 0.38 |
|  | 35-39 years | 134.372 | 50 | 0.37 | 0.28, 0.49 | 0.25 | 0.18, 0.34 |
|  | 40-44 years | 100.548 | 31 | 0.31 | 0.21, 0.44 | 0.21 | 0.14, 0.30 |
|  | 45-49 years | 72.877 | 15 | 0.21 | 0.12, 0.34 | 0.14 | 0.08, 0.23 |
| **Socioeconomic group** | Self-employed | 41.498 | 17 | 0.41 | 0.24, 0.66 | reference | reference |
|  | Upper-level employees | 143.872 | 17 | 0.12 | 0.07, 0.19 | 0.29 | 0.15, 0.57 |
|  | Lower-level employees | 318.356 | 111 | 0.29 | 0.24, 0.35 | 0.71 | 0.43, 1.18 |
|  | Manual workers | 174.397 | 106 | 0.61 | 0.50, 0.74 | 1.48 | 0.89, 2.48 |
|  | Students | 240.155 | 261 | 1.09 | 0.96, 1.23 | 2.65 | 1.62, 4.33 |
|  | Pensioners | 20.526 | 65 | 3.17 | 2.44, 4.04 | 7.73 | 4.53, 13.18 |
|  | Others | 101.734 | 115 | 1.13 | 0.93, 1.36 | 2.76 | 1.66, 4.59 |
|  | Unknown | 70.936 | 68 | 0.96 | 0.74, 1.22 | 2.34 | 1.38, 3.98 |
| **Education** | Upper secondary | 544.173 | 300 | 0.55 | 0.49, 0.62 | reference | reference |
|  | Post-secondary non-tertiary | 7.483 | <5 | 1.13 | 0.003, 0.75 | 0.24 | 0.03, 1.73 |
|  | Short-cycle tertiary | 30.799 | 6 | 0.20 | 0.07, 0.42 | 0.35 | 0.16, 0.79 |
|  | Bachelor | 224.233 | 27 | 0.12 | 0.08, 0.18 | 0.22 | 0.15, 0.32 |
|  | Master | 124.133 | 14 | 0.11 | 0.06, 0.19 | 0.21 | 0.12, 0.35 |
|  | Doctoral | 6.529 | <5 | 0.15 | 0.004, 0.85 | 0.28 | 0.04, 1.98 |
|  | Unknown | 237.125 | 411 | 1.73 | 1.57, 1.91 | 3.14 | 2.71, 3.65 |
| **Marital status** | Unmarried | 805.475 | 623 | 0.77 | 0.71, 0.84 | reference | reference |
|  | Married | 302.036 | 75 | 0.25 | 0.20, 0.31 | 0.32 | 0.25, 0.41 |
|  | Divorced | 63.222 | 60 | 0.95 | 0.72, 1.22 | 1.23 | 0.94, 1.60 |
|  | Widowed | 2.184 | <5 | 0.46 | 0.01, 2.55 | 0.59 | 0.08, 4.21 |
|  | Other | 1.558 | <5 | 0.64 | 0.02, 3.58 | 0.83 | 0.12, 5.90 |

**Table S6. Basic characteristics of the nested case-control study of attempted suicides, only Care Register for Health Care cases included.**

|  | **Cases**  **(N=760)** | | **Controls**  **(N=3040)** | |  |
| --- | --- | --- | --- | --- | --- |
|  | **N** | **%** | **N** | **%** | **p-value** |
| **Marital status** |  |  |  |  | <0.001 |
| Unmarried | 623 | 82.0 | 2460 | 80.9 |  |
| Married | 75 | 9.9 | 473 | 15.6 |  |
| Divorced | 60 | 7.9 | 97 | 3.2 |  |
| Widowed | <5 | NA | 7 | 0.2 |  |
| Other | <5 | NA | <5 | NA |  |
| **Socioeconomic group** |  |  |  |  | <0.001 |
| Self-employed | 17 | 2.2 | 76 | 2.5 |  |
| Upper-level employees | 17 | 2.2 | 259 | 8.5 |  |
| Lower-level employees | 111 | 14.6 | 773 | 25.4 |  |
| Manual workers | 106 | 13.9 | 444 | 14.6 |  |
| Students | 261 | 34.3 | 1032 | 33.9 |  |
| Pensioners | 65 | 8.6 | 47 | 1.5 |  |
| Others | 115 | 15.1 | 224 | 7.4 |  |
| Unknown | 68 | 8.9 | 185 | 6.1 |  |
| **Education** |  |  |  |  | <0.001 |
| Upper secondary | 300 | 39.5 | 1394 | 45.9 |  |
| Post-secondary non-tertiary | <5 | NA | 15 | 0.5 |  |
| Short-cycle tertiary | 6 | 0.8 | 28 | 0.9 |  |
| Bachelor | 27 | 3.6 | 390 | 12.8 |  |
| Master | 14 | 1.8 | 200 | 6.6 |  |
| Doctoral | <5 | NA | 9 | 0.3 |  |
| Missing (including, e.g., missing information on  education other than of primary school level,  school dropouts) | 411 | 54.1 | 1004 | 33.0 |  |
| **Age group** |  |  |  |  | 1.000 |
| 15-19 years | 229 | 30.1 | 916 | 30.1 |  |
| 20-24 years | 224 | 29.5 | 896 | 29.5 |  |
| 25-29 years | 135 | 17.8 | 540 | 17.8 |  |
| 30-34 years | 76 | 10.0 | 304 | 10.0 |  |
| 35-39 years | 50 | 6.6 | 200 | 6.6 |  |
| 40-44 years | 31 | 4.1 | 124 | 4.1 |  |
| 45-49 years | 15 | 2.0 | 60 | 2.0 |  |
| **Severe psychiatric disorder at baseline** | 95 | 12.5 | 27 | 0.9 | <0.001 |
| **Chronic somatic diseases at baseline^**^** | 54 | 7.1 | 133 | 4.4 | 0.003 |
| **HC use in 2017** | 320 | 42.1 | 1530 | 50.3 | <0.001 |
| **Use of psychotropic medication** |  |  |  |  |  |
| antipsychotics | 265 | 34.9 | 58 | 1.9 | <0.001 |
| anxiolytics | 177 | 23.3 | 25 | 0.8 | <0.001 |
| hypnotics and sedatives | 107 | 14.1 | 44 | 1.4 | <0.001 |
| antidepressants | 372 | 48.9 | 202 | 6.6 | <0.001 |
| psychostimulants | 13 | 1.7 | 12 | 0.4 | <0.001 |
| psycholeptics and psychoanaleptics in combination | 5 | 0.7 | <5 | NA | 0.024 |
| **Recent delivery** |  |  |  |  | 0.002 |
| No | 741 | 97.5 | 2867 | 94.3 |  |
| In the previous 6 months | < 5 | NA | 23 | 0.8 |  |
| 6 to 24 months before | 17 | 2.2 | 150 | 4.9 |  |
| **Recent hospitalization due to psychiatric disorder** |  |  |  |  | <0.001 |
| No | 170 | 22.4 | 2741 | 90.2 |  |
| In the previous 6 months | 490 | 64.5 | 166 | 5.5 |  |
| 6 to 24 months before | 109 | 13.2 | 133 | 4.4 |  |

^*^diabetes, multiple sclerosis, epilepsy, connective tissue diseases, ulcerative colitis or Crohn’s disease

HC, hormonal contraception; NA, not available.

**Table S7. Hormonal contraception use in the nested case-control study of attempted suicides. Care Register for Health Care data only.**

|  | **Cases**  **(N=760)** | | **Controls**  **(N=3040)** | |  |
| --- | --- | --- | --- | --- | --- |
|  | **N** | **%** | **N** | **%** | **p-value** |
| **HC group** |  |  |  |  | <0.001 |
| No HC | 644 | 84.7 | 2335 | 76.8 |  |
| Current HC | 116 | 15.3 | 705 | 23.2 |  |
| Combined hormonal contraceptives | 72 | 9.5 | 558 | 18.4 |  |
| Ethinyl-estradiol containing | 54 | 7.1 | 416 | 13.7 |  |
| Estradiol containing | 18 | 2.4 | 142 | 4.7 |  |
| Progestin-only | 44 | 5.8 | 147 | 4.8 |  |
| **Combined hormonal contraceptives** |  |  |  |  |  |
| G03AA07 (levonorgestrel and ethinylestradiol) | <5 | NA | 14 | 0.5 | 0.124 |
| G03AA09 (desogestrel and ethinylestradiol) | 7 | 0.9 | 47 | 1.5 | 0.258 |
| G03AA10 (gestodene and ethinylestradiol) | 11 | 1.4 | 83 | 2.7 | 0.057 |
| G03AA12 (drospirenone and ethinylestradiol) | 26 | 3.4 | 195 | 6.4 | 0.002 |
| G03AA13 (norelgestromin and ethinylestradiol patch) | <5 | NA | 7 | 0.2 | 0.929 |
| G03AA14 (nomegestrol and estradiol) | <5 | NA | 41 | 1.3 | 0.092 |
| G03AA16 (dienogest and ethinylestradiol) | <5 | NA | 11 | 0.4 | 0.515 |
| G03AB08 (dienogest and estradiol-valerate) | <5 | NA | 16 | 0.5 | 0.248 |
| G02BB01 (etonogestrel and ethinylestradiol vaginal ring) | 8 | 1.1 | 59 | 1.9 | 0.131 |
| **Progestin-only oral contraceptives** |  |  |  |  |  |
| G03AC01 (norethisterone) | 5 | 0.7 | 8 | 0.3 | 0.187 |
| G03AC09 (desogestrel) | 38 | 5.0 | 136 | 4.5 | 0.600 |
| **Antiandrogen and estrogen** |  |  |  |  |  |
| G03HB01 (cyproterone and estrogen) | 13 | 1.7 | 86 | 2.8 | 0.109 |

HC, hormonal contraception; NA, not available
